# Virologic characterization of symptom rebound following nirmatrelvir-ritonavir treatment for COVID-19

**DOI:** 10.1101/2022.05.24.22275326

**Authors:** Julie Boucau, Rockib Uddin, Caitlin Marino, James Regan, James P. Flynn, Manish C. Choudhary, Geoffrey Chen, Ashley M. Stuckwisch, Josh Mathews, May Y. Liew, Arshdeep Singh, Zahra Reynolds, Surabhi L. Iyer, Grace C. Chamberlin, Tammy D. Vyas, Jatin M. Vyas, Sarah E. Turbett, Jonathan Z. Li, Jacob E. Lemieux, Amy K. Barczak, Mark J. Siedner

## Abstract

We enrolled seven individuals with recurrent symptoms following nirmatrelvir-ritonavir treatment. High viral loads (median 6.1 log_10_ copies/mL) were detected at enrollment and for a median of 17 days after initial diagnosis. Three of seven had culturable virus for up to 16 days after initial diagnosis. No known resistance-associated mutations were identified.

## Background

Nirmatrelvir-ritonavir, which inhibits the main viral protease of SARS-CoV-2, has been shown to reduce hospitalization in high-risk patients with early stage, symptomatic COVID-19 infection [1]. The US Food and Drug Administration granted emergency use authorization (EUA) status for its use in December, 2021 and it is currently a preferred therapy for ambulatory individuals with COVID-19 at high risk of severe disease [2]. As nirmatrelvir-ritonavir has entered into broad clinical use, reports have emerged of recurrent symptoms in a subset of treated individuals who had initial symptomatic improvement [3]. However, the mechanism and viral characteristics of symptomatic relapse after nirmatrelvir-ritonavir therapy remain unclear. We sought to characterize the virology of symptomatic rebound after nirmatrelvir-ritonavir with longitudinal sampling of individuals with assessment of nasal swabs with viral load quantification, viral culture, and whole genome viral sequencing.

## Methods

### Study participants

The POSITIVES cohort is a longitudinal study of individuals with COVID-19 infection that aims to characterize virologic and immunologic aspects of infection [4–6]. To better understand the clinical phenomenon of symptom relapse after nirmatrelvir-ritonavir treatment, we selectively enrolled seven ambulatory individuals recently treated with nirmatrelvir-ritonavir with recurrent symptoms after initial resolution. Individuals were identified through the Mass General Brigham health system or upon referral from treating providers and contacted by phone for informed consent at the time of symptom recurrence. Following enrollment, anterior nasal (AN) swabs were collected and placed into viral transport media three days per week until PCR negativity. Swabs were simultaneously analyzed for viral RNA level by qRT-PCR, viral whole genome sequencing, semiquantitative viral culture, and laboratory-based rapid antigen testing.

### Viral load quantification

Viral transport media was centrifuged for 2 hours at 21,000 x g and 4°C to pellet virions. TRIzol-LS™ Reagent (ThermoFisher) was then added to the pellets, and samples were subsequently incubated on ice for 10 minutes. Chloroform (MilliporeSigma) was added to each sample, and the resulting mixtures were then vortexed and centrifuged for 15 minutes at 21,000 x g and 4°C. The clear aqueous layer was collected and concentrated using isopropanol precipitation. RNA was washed with cold 70% ethanol before being resuspended in DEPC-treated water (ThermoFisher). SARS-CoV-2 viral RNA was tested with a qPCR assay using the US CDC 2019 nCoV_N1 primer and probe set and quantified using a standard curve.

### SARS-CoV-2 culture

Viral culture was performed in the BSL3 laboratory of the Ragon Institute of MGH, MIT, and Harvard. Viral culture was assessed semi-quantitatively by median tissue culture infectious dose assay (TCID50) as previously reported [4,6]. Briefly, aliquoted viral transport media were filtered with 0.45-0.65um centrifugal filters and added to Vero-E6 cells plated in DMEM culture media supplemented with 2% fetal bovine serum, HEPES, antibiotic-antimycotic solution and 5ug/mL of polybrene. Each sample was added to the cells in four replicates and serially diluted six times in 5-fold increments in 96-well format. The infection was performed by spinfection for one hour at 2,000x g at 37°C. The cytopathic effect (CPE) was scored seven days post-infection with light microscopy and TCID50/mL titers were calculated using the Reed-Muench method. The supernatant of wells showing CPE was harvested for RNA extraction and viral sequence confirmation.

### SARS-CoV-2 Whole Genome Sequencing

We sequenced SARS-CoV-2 genomes using the Illumina COVIDSeq Test protocol as previously described [4]. We constructed sequencing libraries using the Illumina Nextera XT Library Prep Kit and sequenced them using an Illumina NextSeq 2000 instrument. Complete genomes (>24,000 assembled base pair length) were assigned a Pango lineage using pangolin v4.0.6. All samples were deposited to GenBank (Bioproject Accession PRJNA759255) and GISAID. Notable amino acid mutations and sequence quality were analyzed using Nextclade v1.14.1.

### Antigen testing using Abbott BinaxNow SARS-CoV-2 Rapid Antigen Assay

Frozen viral transport media aliquots were thawed on ice and 50*u*L was transferred to a test tube. Swabs from BinaxNow antigen kits (Abbott, Chicago, IL) were immersed into the liquid until fully absorbed and then tested according to manufacturer’s instructions as previously described [7]. After 15 minutes, result were interpreted as positive, negative, or discordant by a reader blinded to the viral load result.

### Ethical considerations

Study procedures were approved by the human subject’s review committee at Mass General Brigham and all participants gave informed consent to participate.

## Results

Participant demographic details are provided in Supplemental Table 1. All seven participants reported symptom improvement and conversion to negative home-based antigen testing following treatment with nirmatrelvir-ritonavir. Symptoms recurred a median of 9 days after initial positive test or 4 days after completion of the nirmatrelvir-ritonavir course. At study enrollment, all seven participants had a detectable viral load (median 6.1 log_10_ copies per mL [range 4.2-7.3]). A detectable viral load was identified for a median of 17 days (range 14-20) after their initial PCR test and for 12 days (range 9-16) after initiation of nirmatrelvir-ritonavir (**Fig. 1**). Enrollment sample viral cultures were positive in three of seven individuals. In these three individuals, cultures were positive up until 10,16, and 16 days, respectively from the time of their initial PCR test and 5, 11, and 11 days after completion of the course of nirmatrelvir-ritonavir.

**Figure 1.**
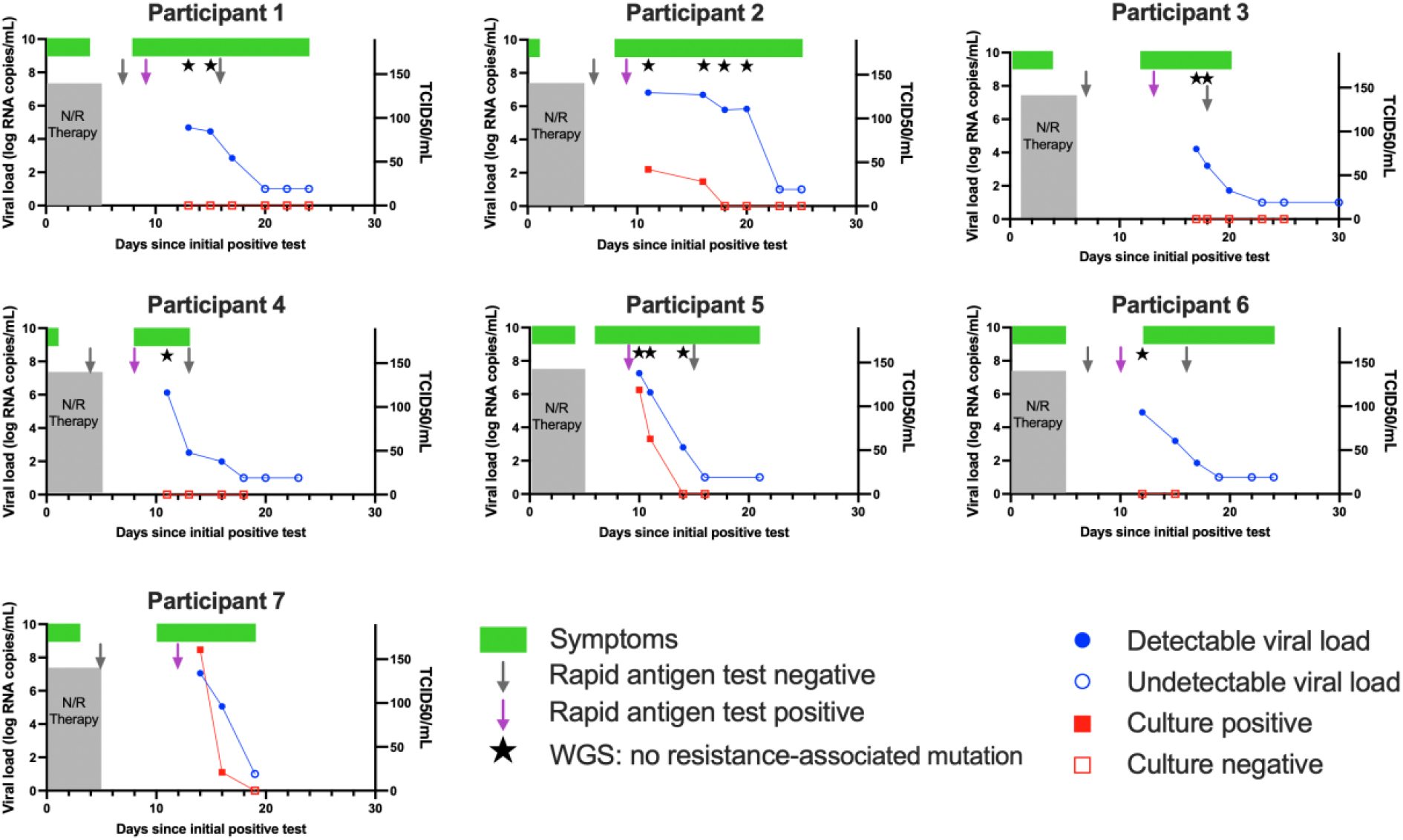
Virologic and clinical course of individuals with symptomatic recurrence of COVID-19 following nirmatrelvir-ritonavir treatment. WGS: whole genome sequencing, N/R: nirmatrelvir-ritonavir

We sequenced virus in six of the seven participants after completion of therapy. We found no known resistance-associated mutations in nsp5 encoding the main SARS-CoV-2 protease (Supplemental Figure 1) or in any of the protease cleavage sites.

Finally, using laboratory-based antigen testing, we found high concordance between antigen and viral culture testing (92%, 24/26). Although there were two specimens that were antigen positive and culture negative, no specimens were antigen negative and culture positive.

## Discussion

Symptomatic relapse after nirmatrelvir-ritonavir therapy for early stage COVID-19 infection is associated with high viral load and, in a subset of individuals, culturable virus. We identified live virus at up to 11 days after completion of nirmatrelvir-ritonavir therapy (16 days from the pre-treatment PCR test). By contrast, we recently reported that untreated outpatients infected with the Omicron variant SARS-CoV-2 shed viable virus for a median of 5 days after an initial positive test [6]. These data reinforce the importance of testing and isolation guidelines for individuals with recurrent symptoms after nirmatrelvir-ritonavir treatment, irrespective of intermediate negative antigen testing or initial symptom resolution. Because live viral shedding can occur at the time of relapse, restarting monitoring and isolation from the time of relapse may be warranted.

Although field-based tested is needed to confirm our findings, we found high concordance between laboratory-based rapid antigen testing and culture positivity, with no specimens that were antigen negative and culture positive. Consequently, antigen test-based monitoring of individuals with relapse after therapy holds promise as a means of signaling a safe release from isolation.

Finally, we did not identify emergence of any resistance-associated polymorphisms in any of the six specimens that were sequenced. Our findings add support to others that drug resistance does not appear to be a significant contributor to relapse [3,8] and suggest that nirmatrelvir-ritonavir may retain activity in most cases of symptom recrudescence.

Our study should be interpreted in the context of limitations. Most notably, this represents a small case series so precise estimates of culture positivity, duration of viral shedding, or incidence of drug resistance cannot be made. We also enrolled individuals after clinical rebound, so cannot determine the incidence of this syndrome among individuals taking nirmatrelvir-ritonavir treatment. Finally, we use viral culture as a proxy measure of contagiousness but cannot quantify the risk of transmission for this patient population.

In summary, recurrent clinical disease after nirmatrelvir-ritonavir therapy for COVID-19 is associated with high viral load and, in some cases, culturable virus. Culturable virus was present for up to two weeks after completion of therapy. Consideration should be given to revising public health guidelines to specifically recommend repeat testing and isolation in these cases. Future work is needed to better understand the causes, clinical significance, and public health consequences of symptomatic relapse after nirmatrelvir-ritonavir.

## Data Availability

All data produced in the present study are available upon reasonable request to the authors

## Acknowledgements

The authors would like to thank Vamsi Thiriveedhi, Ha Eun Cho, and Seamus Carroll for assistance with sequencing. We would also like to thank the participants for their time and participation, as well as the many physicians for referral of patients to our study.

