## Supplement for "Virologic characterization of symptom rebound following nirmatrelvir-ritonavir treatment for COVID-19"

**Supplemental Table 1**. Cohort Characteristics

| ID | Gender | Age Range | Completed COVID-19 Vaccinations at Enrollment | Days from End of N/R Therapy to Symptom Relapse | Days from End of N/R Therapy to Antigen Positivity | Days from End of N/R Therapy to Study Enrollment | Enrollment Viral Load (Log10 copies/mL) | Days from Diagnosis to Negative PCR | Positive Culture After Relapse (Y/N) | Days from Diagnosis to Negative Viral Culture |
|---|---|---|---|---|---|---|---|---|---|---|
| 1 | F | 31-35 | 4 | 4 | 5 | 9 | 4.7 | 20 | N | N/A |
| 2 | M | 51-55 | 3 | 5 | 5 | 8 | 6.8 | 23 | Y | 16 |
| 3 | F | 46-50 | 3 | 7 | 4 | 7 | 4.2 | 23 | N | N/A |
| 4 | F | 51-55 | 5 | 4 | 4 | 7 | 6.1 | 18 | N | N/A |
| 5 | F | 46-50 | 4 | 2 | 5 | 6 | 7.3 | 16 | Y | 14 |
| 6 | F | 61-65 | 3 | 3 | 6 | 8 | 4.9 | 19 | N | N/A |
| 7 | F | 36-40 | 3 | 6 | 8 | 10 | 7.1 | 19 | Y | 19 |

N/R: nirmatrelvir-ritonavir
N/A: not applicable

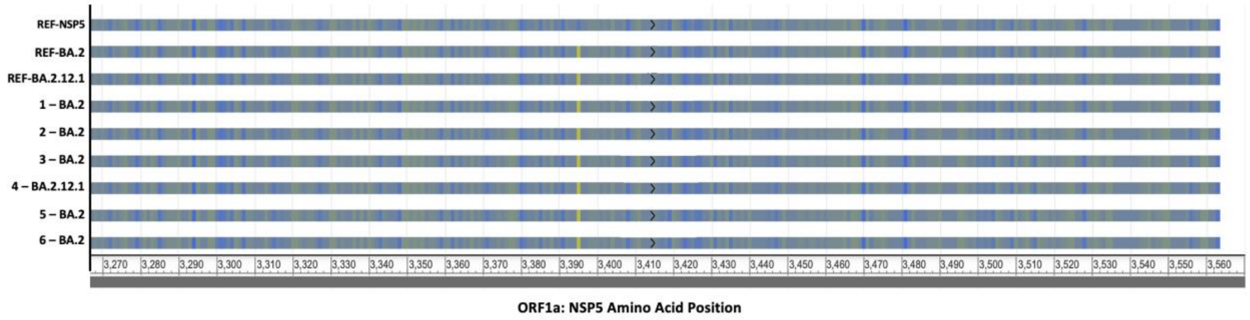


**Supplemental Figure 1.** Amino acid sequence of NSP5 (main viral protease, or Mpro) for individuals with recurring infection following nirmatrelvir-ritonavir treatment, compared to reference WT genome (GenBank YP_009725301.1). Ref-BA.2 (USA/MA-MGB-06410/2022) and Ref-BA.2.12.1 (USA/MA-MGB-05617/2022) genomes were collected via MGH surveillance sequencing and used as lineage-specific references. No mutations were found in drug-resistance conferring regions of the NSP5 gene of the six sequenced specimens.
